# Cervical lymph node B cell subset resists depletion and sustains lytic Epstein Barr-virus signature in patients with multiple sclerosis

**DOI:** 10.64898/2026.10.25.26366311

**Authors:** Yigit K. Babal, Joona Sarkkinen, Ella Ahvenainen, Annika Susi, Mika Saarela, Goran Kurdo, Sini M. Laakso

## Abstract

B cells have arisen as key in multiple sclerosis (MS) pathogenesis by the high efficacy of B cell depleting CD20 monoclonal antibodies and by the identification of Epstein-Barr virus (EBV) as a prerequisite factor. We previously demonstrated an expansion of memory and double negative (DN) B cells in the deep cervical lymph nodes (dcLNs) of newly diagnosed patients with MS (pwMS), with DN B cells showing a molecular signature consistent with lytic EBV infection. Effect of anti-CD20 therapies for pwMS require sustained administration, which led us to hyhenhesize that incomplete effects on the dcLNs, a natural site for lymphocyte priming towards the central nervous system (CNS), might be sustaining the pathology. Using single-cell transcriptomics and paired sampling, we characterized the effects of anti-CD20 therapies on dcLNs compared to circulation and to other therapies. We observed switched (SM) and unswitched (USM) memory B cells to be depleted uniquely by anti-CD20 in both dcLNs and circulation, whereas DN B cells of the dcLNs were not. Moreover, EBV lytic signature was retained in DN B cells during anti-CD20, and using clonal trajectory we saw that the DN B cells continued to contribute to the plasmablast differentiation within dcLNs. EBV targeting of expanded memory CD8 T cells, as well as CD8 T cell exhaustion, were retained during anti-CD20 therapy within the dcLNs. Our findings suggest a sustained, EBV-driven process in pwMS during anti-CD20 in the CNS-targeting priming dcLNs, with therapeutic implications to be investigated further.

**One Sentence Summary:** Cervical lymph nodes show sustained, EBV-driven pathology in multiple sclerosis during anti-CD20 therapy unseen in peripheral blood.

## INTRODUCTION

Multiple sclerosis (MS) is a chronic demyelinating disease of the central nervous system (CNS) characterized by influx of T and B cells from the periphery (*1*). High efficacy of B cell depleting therapies has highlighted T and B cell interactions in the disease mechanism (*2–5*). A B cell resident, namely Epstein–Barr virus (EBV), has been identified as a prerequisite for MS (*6*), and abnormal anti-EBV T cell responses (*7*) and unstable EBV latency (*8, 9*) characterize this lifelong viral persistence in patients with MS (pwMS), which typically takes place in secondary lymphoid organs such as the deep cervical lymph nodes (dcLNs) (*10*).

The dcLNs are strategically positioned as a critical interface between the CNS and the peripheral immune system by draining CNS antigens through the meningeal lymphatic vessels and the associated glymphatic system (*11*). In post-mortem samples of pwMS, the lymphocytes in the demyelinating lesions have been shown to derive from the dcLNs, suggesting CNS-targeting is programmed in the dcLNs in pwMS (*12*). Enlargement of dcLNs in MS was suggested by an ultrasound study (*13*), and a case report saw marked lymphoblasts within the dcLN in a postmortem setting (*14*). We previously demonstrated an expansion of memory and IgD and CD27 double negative (DN) B cells in the dcLNs of newly diagnosed pwMS compared to healthy controls, accompanied by molecular signatures consistent with lytic EBV infection in the DN B cells (*15*). We also previously reported T follicular helper (Tfh) cells to be increased in MS lymph nodes (LNs) even during the prediagnostic phase (*16*).

The effects of disease-modifying therapies (DMTs) across distinct compartments in MS remain incompletely understood. Most studies have focused on circulating cells, but interaction of immune cells takes place primarily within lymphoid tissues (*12, 17*). It is unknown whether the efficacy of DMTs is a result of the observed effects on circulating cells or whether reaching lymphatic organs is needed. In the case of macromolecules such as the monoclonal antibodies binding CD20, CNS is not reached across an intact blood-brain barrier, but dcLN engagement has been suggested (*18*). The magnitude of this effect is still unresolved. Addressing this gap is essential to understanding the origins and persistence of immune dysregulation in MS.

Effect of anti-CD20 therapies for pwMS require sustained administration (*19, 20*), which led us to hypothesize that incomplete depletion of the dcLN B cell compartment might be sustaining the pathology. Using single-cell transcriptomics and sequential sampling, we further characterize the B cell compartment of pwMS at diagnosis and the effects of anti-CD20 treatment (rituximab and ofatumumab) in the dcLNs compared to the circulation. We explore whether B cell depletion changes the EBV-targeting CD8 T cell responses within the dcLNs. To understand the specificity of the findings to anti-CD20 therapy, we analyze paired dcLN and peripheral blood samples from pwMS before and during treatment with dimethyl fumarate (DMF) and natalizumab (NTZ).

## RESULTS

### DN B cells of the dcLNs are not fully depleted during anti-CD20 therapy

To investigate the effect of anti-CD20 on dcLNs and to compare to the effects on PBMC and to those of other DMTs, 5’ scRNAseq along with VDJ sequencing was performed for 13 pwMS sampled at the diagnostic phase before DMT initiation and eight pwMS during a DMT, of which five on anti-CD20 (three on rituximab, two on ofatumumab), two on dimethyl fumarate (DMF) and one on natalizumab (NTZ), and compared also to data on three healthy controls. The dcLN scRNAseq data on the three healthy controls and six pwMS at the diagnostic phase are presented in a previous publication (*15*). Of the eight pwMS included during a DMT, seven had sampling at the diagnostic phase as well and thus represent a sequential analysis. Sequential PBMC scRNAseq was available for four pwMS.

After excluding low-quality cells, performing sample integration and cell type annotation, scRNAseq data of a total of 225,733 dcLN cells and 84,385 PBMC were taken for further analysis. We performed multi-layered cell type clustering and annotation using automatic annotation with SingleR (with human primary cell atlas, and Monaco Immune reference) and manual curation (Figure 1A-C). Manual annotation was carried out with known marker gene expressions from literature, publicly available marker databases and our previous study (*15*)(Figure 1D-E, Figure S1). Total number of cells per sample and sequencing protocol can highly affect the statistical analysis of cell-type frequency distributions. Therefore, we applied a sum-constrained beta-binomial modelling approach to account for these effects when comparing cell proportions across DMTs (*21*).

**Fig. 1:**
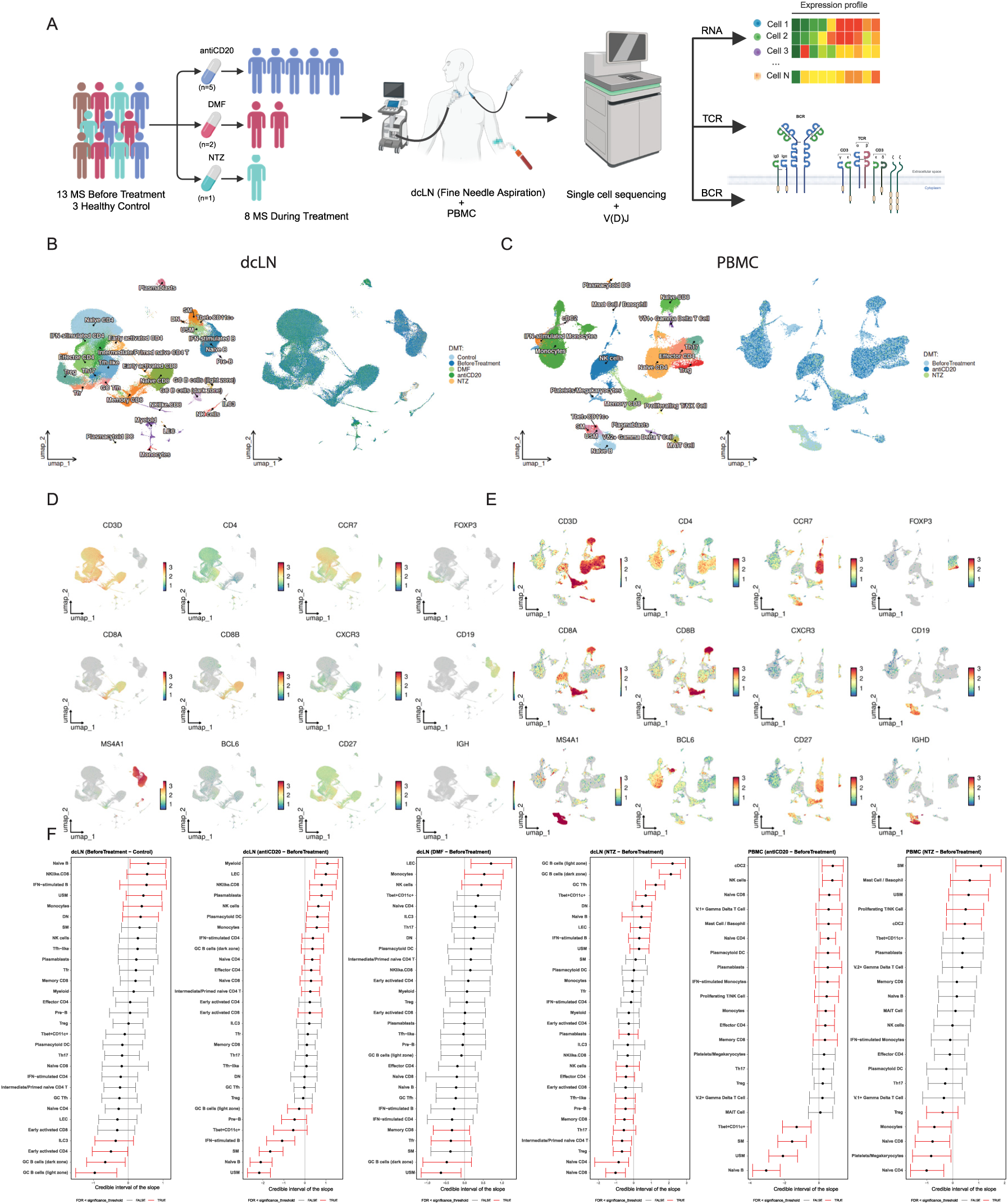
Immune landscape of dcLNs and circulation in pwMS under DMTs. (**A**) Methodological illustration of the study. (**B-C**) Uniform Manifold Approximation and Projection (UMAP) plot of identified cell clusters and sample distribution of (**B**) dcLNs and (**C**) blood. (**D-E**) UMAP plots of key marker gene expression of major T and B cell clusters in (**D**) dcLN and (**E**) blood. (**F**) 1D interval plots of sum-constrained beta-binomial model for proportional effect of DMTs in dcLNs and PBMC in pwMS.

Consistent with our earlier findings, naïve and memory B cells were significantly increased in the dcLNs of pwMS prior to DMT initiation at the diagnosis compared to healthy controls. During anti-CD20 therapy we saw a significant depletion of the B cell compartment, including naïve and memory B cells in both the dcLNs and PBMC. DMF showed milder effects, with significant depletion of memory B cells but not of naïve B cells (Figure 1F). Unlike the depletion therapies, NTZ seemed to increase the proportion of memory B cells in the dcLNs.

In the T cell compartment, we observed significantly elevated NK like-CD8 T cells in dcLNs of pwMS at the diagnostic phase compared to healthy controls. During anti-CD20, we saw increases in naïve CD4/CD8, intermediate and early activated CD4, effector CD4 and IFN-stimulated CD4 T cells in dcLNs, whereas in the PBMC, naïve CD4/CD8, effector CD4 and memory CD8 T cells were increased. During DMF, T follicular regulatory (Tfr) and memory CD8 T cell subsets decreased in the dcLNs. NTZ effected oppositely the T cell compartment compared to B cells, as naïve CD4/CD8 and T regulatory cells (Treg) were significantly decreased in both dcLNs and PBMC (Figure 1F).

To investigate the effect of anti-CD20 on the B cell compartment in detail, we sub-clustered B cells from both the dcLNs and PBMC. Memory B cell clusters were classified as switched (SM) and unswitched (USM) based on IGHD expression (*22*). A Tbet+CD11c+ B cell cluster representing activated atypical B cells prevalent in autoimmune diseases known as type 2 double-negative (DN) B cells was also identified in both the dcLNs and PBMC (*22*). A more generic DN B cell cluster was further identified by the absence of IGHD and CD27 expression in the dcLNs, which was not detected in PBMC. Specific to the dcLNs, we identified an interferon-stimulated (IFN) B cell cluster characterized by significant expression of two canonical type I IFN-stimulated genes IFI44L and IFIT1 encoding antiviral effector proteins that restrict viral replication and modulate innate immune signaling downstream of the JAK-STAT pathway (*23–25*) that were exclusively expressed by this cluster (Figure 2A–D).

**Fig. 2:**
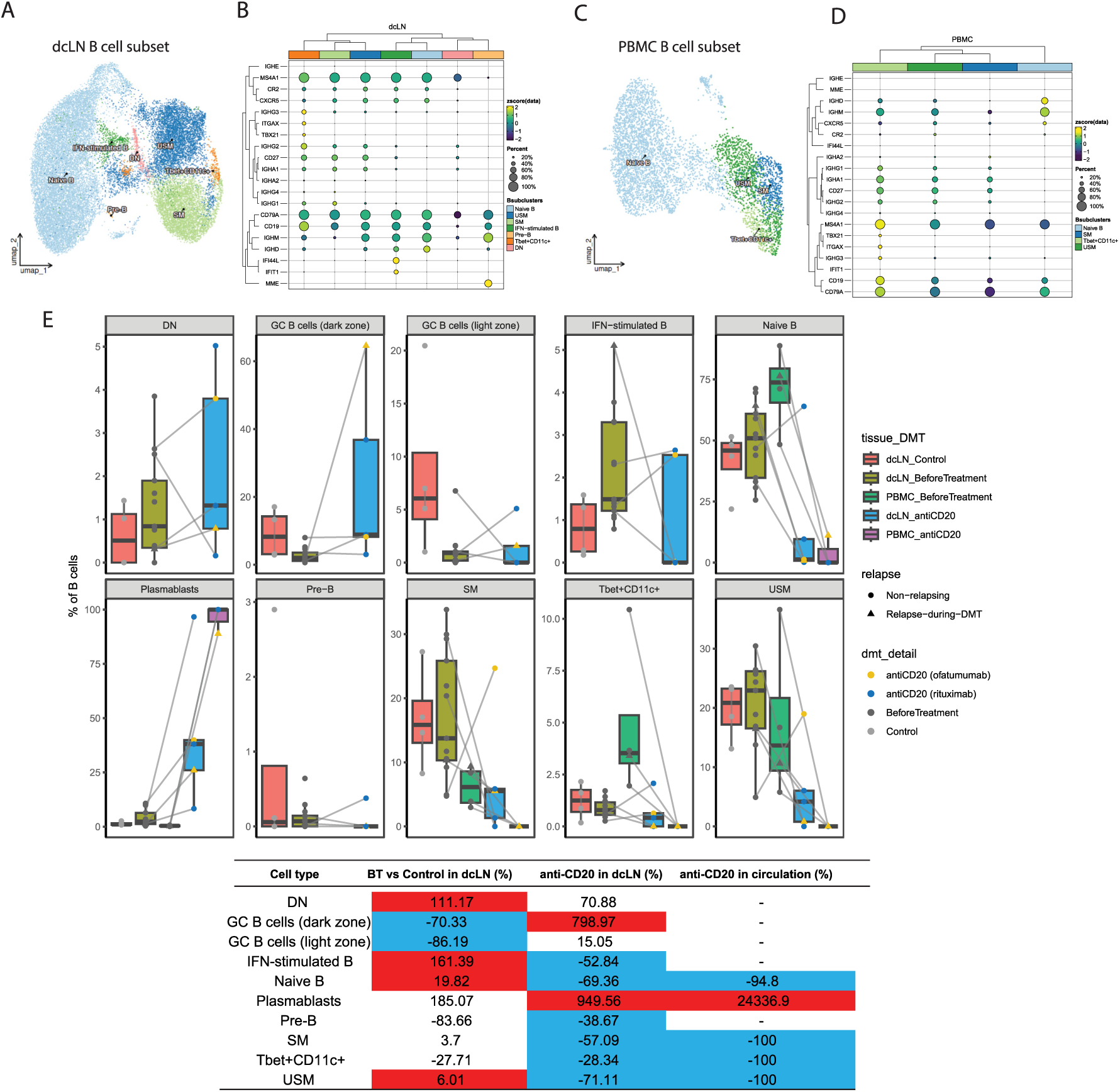
DN B cells are increased in MS dcLNs and resist anti-CD20 therapy. (**A-D**) UMAP and dot plots of subclustered B cell compartment in (**A-B**) dcLNs and (**C-D**) circulation. Dot plots present the expression of key marker gene expression used to identify B cell subsets in (**B**) dcLNs and (**D**) circulation. (**E**) Box plots showing percentage of B cell subgroups in dcLNs and circulation. Table on the bottom shows relative percentage change of B cell percentages in comparison of pwMS before treatment (BT) to healthy controls (Control) in the dcLNs, anti-CD20-treated pwMS to before treatment in dcLNs, and anti-CD20-treated pwMS to before treatment in circulation. Red (increased) and blue (decreased) indicate significant change by proportional change model.

In dcLNs of pwMS at diagnosis compared to healthy controls, naïve B cells were increased by 19.82%, USM by 6.01%, IFN-stimulated B cells by 161.39%, and DN B cells by 111.17%. During anti-CD20 therapy, of PBMC, the SM, USM and Tbet+CD11c+ B cells were fully (-100%) and naïve B cells (-94.8%) almost fully depleted. For the dcLNs of pwMS during anti-CD20, naïve B cells (-69.36%), SM (-57.09%), USM (-71.11%), and IFN-stimulated B cells (-52.84%) were all significantly depleted, whereas Tbet+CD11c+ B cells (-28.34%) were only modestly reduced and DN B cells were not (+70.88%) (Figure 2E). Notably, DN B cells displayed relatively reduced CD20 expression compared to other B cell subsets (Figure 2B). When comparing to the effects of other therapies, we saw that the DN B cells did not proportionally change during DMF therapy but increased during NTZ (Figure 1F).

### Intranodal T-cell-B-cell interactions change during anti-CD20 therapy

We then performed differential gene expression analysis for each cell type comparing samples taken before treatment to those taken during anti-CD20 therapy of pwMS, combining a pseudo-bulk approach with consensus analysis using three well-known DE analysis tools (DESeq2 (*26*), edgeR (*27*), and limma (*28*)). Genes were identified as DEGs when they reached statistical significance (adjusted p-value < 0.05) in at least two of the three tools. Using these criteria, the majority of DEGs were detected in naïve B cells, with 98 upregulated and 33 downregulated genes in the dcLNs and 135 upregulated and 225 downregulated genes in the PBMC in the comparison between anti-CD20 and before treatment pwMS (Figure S2A-B). The remaining cell types showed very few or no DEGs. When comparing DEGs of naïve B cells across compartments, only 2% of upregulated genes (4 genes) and 4% of downregulated genes (11 genes) were shared between the dcLNs and PBMC (Figure S2C). Although the DEGs were largely compartment-specific, Gene Ontology (GO) over-representation analysis revealed convergent biological processes across both compartments. Specifically, in dcLNs, downregulated genes of naïve B cells were predominantly associated with cellular biosynthesis processes, while upregulated genes were enriched for IFN responses. In PBMC, downregulated genes of these cells were enriched for MHC protein complex related processes and upregulated genes were enriched for cellular metabolism (Figure S2D-E). Collectively, these results suggest that surviving naïve B cells during anti-CD20 therapy exhibit a transcriptional signature consistent with elevated cellular stress and possible cell death.

As a complementary approach to the differential gene expression analysis, we characterized B cell–T cell interactions during anti-CD20 therapy and performed ligand-receptor-based cell–to-cell communication analysis using CellChat (*29*) in both dcLNs and PBMC. As expected, the near-complete depletion of B cells in peripheral blood under anti-CD20 prevents the detection of substantial B–T cell interactions in PBMC. In contrast, dcLN analysis revealed distinct ligand-receptor communication networks between DN B cells, IFN-stimulated B cells, and Tbet⁺CD11c⁺ B cells with early activated CD4⁺, effector CD4⁺, germinal center follicular helper (GC Tfh), IFN-stimulated CD4⁺, and early activated CD8⁺ T cells (Figure S2F). Notably, the same three ligand-receptor pairs of galectin-9 (LGALS9), a T cell modulator linked with promoting T cell exhaustion and regulation (*30*), with P4HB, CD45, and CD44 were consistently identified between several CD4+ T cell subsets and all three B cell subsets.

### DN B cells retain EBV lytic signature during anti-CD20 therapy

To understand B cell clonal overlap and survival, we evaluated the BCR repertoire in the dcLNs of pwMS at the diagnostic phase and during anti-CD20 therapy. A total of 1,185 B cells with matched BCR repertoire and single-cell RNA-seq data were included in the repertoire analysis using the scRepertoire package (*31*). Significant differences in Shannon diversity were observed only for DN B cells at the diagnostic phase in pwMS compared to healthy controls, and for naïve B cells during anti-CD20 in pwMS compared to the diagnostic phase before therapy: Both DN and naïve B cells showed reduced Shannon diversity, consistent with clonal expansion (Figure 3A-C, Figure S3A).

**Fig. 3:**
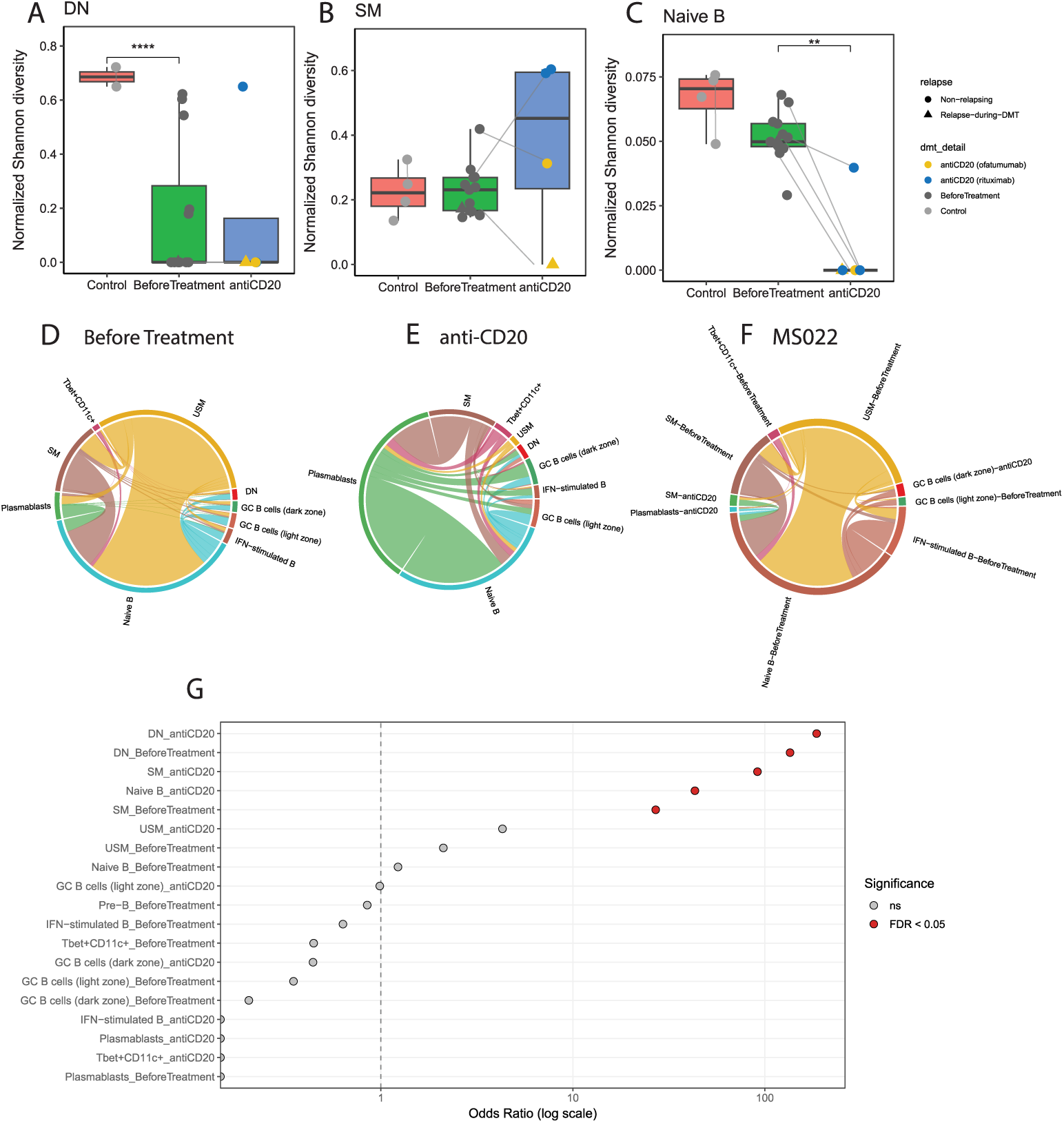
DN B cells show EBV lytic gene signature and share clonality with naïve, memory B cells and plasmabasts in dcLNs of pwMS. (**A-C**) Box plots show BCR diversity of DN, naïve and SM in dcLNs by normalized Shannon diversity measurement. (**D**) Chord diagram shows intra-cellular BCR clonal share of B cell compartment in dcLNs of pwMS before treatment, (**E**) after anti-CD20 therapy and (**F**) for the relapsing patient (MS022). (**G**) Odds ratios (ORs) for the overrepresentation of EBV lytic host transcripts in the B cell subsets analysed as presented by SoRelle et al. (*35*).

To further characterize the potential clonal origins of B cell subsets in the dcLNs, germline B cell clonal lineage trees were constructed using the Immcantation suite (*32–34*). A total of 14 unique clonal trees encompassing all B cell subsets of the dcLNs before treatment and during anti-CD20 therapy were reconstructed (four biggest clones are shown in Figure S4). Here, DN, Tbet+CD11c+ and IFN-stimulated B cells shared clonal relationships along a naïve–memory–plasmablast differentiation trajectory that only seldom involved the GC reaction (Figure 3D–E, Figure S3B). In pwMS at the diagnostic phase, all these B cell subtypes were clonally connected to naïve and USM B cells, whereas during anti-CD20 therapy, these clonal relationships shifted toward naïve B cells and plasmablasts (Figure 3D–E) (*22*).

The patient who relapsed during anti-CD20 therapy and was sampled shortly thereafter (MS022) had lowest Shannon diversities across several B cell subsets suggesting clonal B cell activation (Figure 3A-C). Moreover, GC dark zone B cells showed the most pronounced increase of any subset. Before treatment, the frequency of GC dark zone B cells in this patient was below that of healthy controls (as observed across all pwMS; Figure 2E), but after relapse on anti-CD20 therapy these cells expanded markedly. Importantly, the post-relapse GC dark zone B cells shared BCR clones with naïve, USM, SM and, notably, IFN-stimulated B cells present at diagnosis (Figure 3F). These findings suggest that during relapse on anti-CD20, progeny of pre-treatment IFN-stimulated B cells acquire a GC phenotype and continue to differentiate. By contrast, during more stable disease on anti-CD20, DN, T-bet+CD11c+ and IFN-stimulated B cells appear to rely more on an extrafollicular pathway, in addition to GC reactions, to generate memory B cells or plasmablasts (Figure S4) (*22*).

We then compared the transcriptional signature of EBV host lytic markers of the dcLN B cell subsets to those of lymphoblastoid cell line as described by SoRelle et al. (*35*). Enrichment analysis revealed that in pwMS, DN and SM B cells both at the diagnostic phase and during anti-CD20 therapy, as well as naïve B cells during anti-CD20, showed a significant EBV host lytic gene signature (FDR < 0.05; Fisher’s exact test, odds ratios DN > 100, SM and Naïve B cells > 10; Figure 3G). These results suggest that the clonal relationships observed among surviving DN, SM and naïve B cells in the dcLNs under anti-CD20 therapy are affected by EBV.

### Expanded CD8 T cell repertoire against EBV in the dcLNs is sustained during anti-CD20

We next investigated the T cell receptor (TCR) repertoire of the dcLNs. Repertoire analysis was performed using the scRepertoire package (*31*), and clonal expansion was defined as clonotypes represented by a minimum of two clones per sample. Consistent with our earlier findings, memory CD8 T cells constituted the most clonally expanded population (Figure 4A). Using these expanded TCR clonotypes from memory CD8 T cells, we performed TCR-epitope matching against the public VDJdb repertoire using tcrdist3 (*36*). The proportion of expanded memory CD8 T cells in the dcLNs was similar between pwMS at the diagnosis and during anti-CD20 treatment and the EBV epitope targeting was not affected by anti-CD20 therapy (Figure 4B). NK-like CD8 T cells was the second most expanded clonal population, and we observed very similar EBV targeting by them (highest for EBNA3A, EBNA3B and BZLF1) in both the before treatment and during anti-CD20 taken samples.

**Fig. 4:**
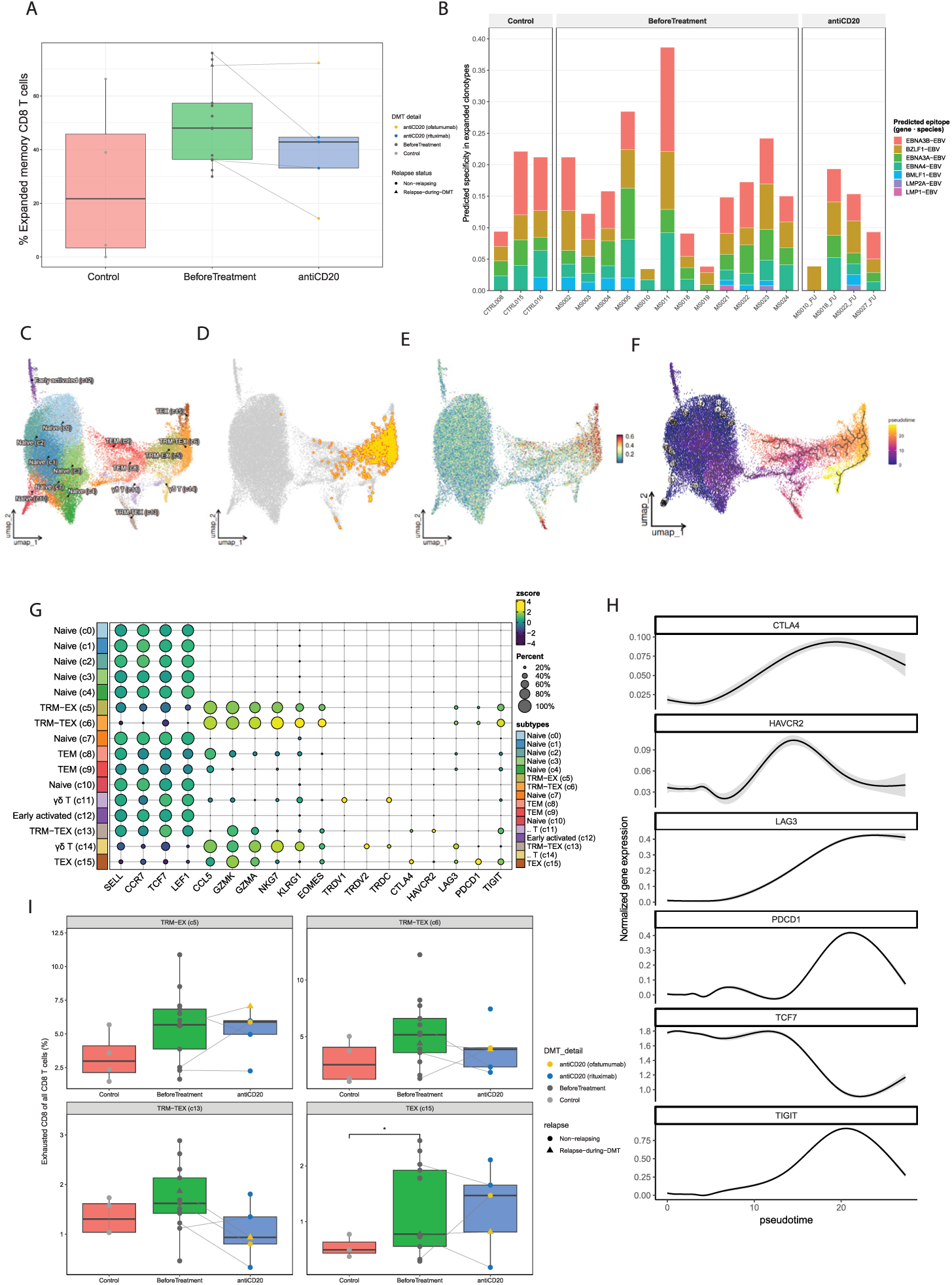
CD8 T cell EBV targeting and exhaustion are sustained during anti-CD20 therapy. (**A**) Box plots presenting percentage of expanded memory CD8 T cells. Paired TCR clonotypes ≥ 2 times within a sample was considered as an expanded clone. (**B**) Stacked bar plot of EBV epitope targeting in expanded CD8 T cell clonotypes. The VDJdb database was referenced for TCRαβ and TCRβ clonotypes using tcrdist3. (**C**) UMAP plot of CD8 T cell subclusters. (**D**) Mapping of expanded T cells over CD8 T cell UMAP. (**E**) UMAP plot showing exhaustion module score. (**F**) Monocle exhaustion trajectory pseudotime represented in UMAP. (**G**) Dot plot showing key marker gene expression including exhaustion markers used in the identification of CD8 T cell subsets. (**H**) Smooth line plots showing significantly changed dynamic exhausted gene expression of CD8 T cells over pseudotime. (**I**) Box plots of exhausted CD8 T cell clusters in dcLNs of pwMS before treatment, during anti-CD20 and of healthy controls.

To address potential CD8 T cell exhaustion, an exhaustion module score was first calculated using a previously described marker gene set comprising TIGIT, HAVCR2, PDCD1, CD244, LAG3, TCF7, and CTLA4 (Figure 4C–E) (*15*). Pseudotime trajectory analysis was then performed using Monocle3 (*37, 38*), with the naïve CD8 T cell cluster designated as the root (Figure 4F). Integrating the exhaustion module scores with trajectory-associated marker gene expression (Figure 4G), we annotated CD8 T cell subclusters and identified three distinct exhausted CD8 T cell subpopulations: tissue-resident exhausted memory T cells (TRM-EX, cluster 5), defined by tissue residency markers LRG1 and EOMES alongside TIGIT and LAG3, tissue-resident terminally exhausted memory T cells (TRM-TEX, clusters 6 and 13), characterized by co-expression of TIGIT, PDCD1 and LAG3, and terminally exhausted memory T cells (TEX, cluster 15), defined by high expression of PDCD1, TIGIT and LAG3 in the absence of KLRG1 and EOMES. Then we mapped expanded CD8 T cells into the sub-clusters, and saw that most of the expanded cells belonged to TRM-EX and TREM-TEX (cluster 6) cells (Figure 4D). Also non-exhausted clusters including effector memory T cells and γδ T cells were identified along the trajectory. Progressive upregulation of exhaustion-associated genes along pseudotime was observed (Figure 4H). TEX cells were significantly elevated in pwMS prior to treatment compared to healthy controls; however, they were not decreased following anti-CD20 therapy (Figure 4I). The exhausted CD8 T cell clusters showed similar ligand-receptor interaction with B cell subsets as other T cell compartments, with interactions especially with DN, IFN-stimulated and Tbet+CD11c+ B cells (Figure S2G). These results suggest that anti-CD20 does not normalize CD8 T cell exhaustion and EBV targeting in pwMS.

## DISCUSSION

By performing a comprehensive single-cell profiling of dcLNs and peripheral blood from pwMS receiving anti-CD20 therapy and comparing the findings to pre-treatment sampling and to effects of other DMTs, we saw that while the majority of B cells were depleted in both circulation and the dcLNs uniquely by anti-CD20, we identified a distinct DN B cell population in the dcLNs that persisted and continued to express a signature of lytic EBV infection. This DN B cell subset maintained clonal connectivity to plasmablasts suggesting ongoing maturation. EBV targeting by expanded CD8 T cell clones and CD8 T cell exhaustion detected at the diagnostic phase were retained during anti-CD20 therapy in the dcLNs. Altogether these findings suggest an ongoing EBV-mediated process in the dcLNs of pwMS.

Effect of anti-CD20 therapies on LNs has been investigated for patients with rheumatoid arthritis, where selective sparing of SM B cells (*39*) or no clearance of intranodal B cells (*40*) was reported. However, the sampling was performed only four weeks after rituximab infusion in these studies, a time frame where at least in pwMS relapses and new lesions are still observed (*41*). For patients with neuromyelitis optica spectrum disorder, especially repeated intravenous rituximab dosing was efficient in clearing the dcLNs of autoantigen-specific memory B cells, suggesting good intranodal effects (*18*). Our study had the median on anti-CD20 therapy time of 13.5 months before follow-up sampling, thus better representing a steady-state effect where most but not all B cell subsets were depleted also from LNs.

The finding of increased DN B cells compared to healthy controls in the dcLNs in our previous study (*15*) remained a constant feature of pwMS here in a larger cohort. These cells were resistant to anti-CD20 mediated depletion and represent a heterogeneous population broadly indicated in autoimmune pathology, expanded for example in systemic lupus erythematosus, rheumatoid arthritis and also reported increased in the circulation and CNS of pwMS (*42–45*). Of special interest in autoimmunity is the Tbet+ subset of DN B cells, also referred to as DN2 or age-associated B cells (ABCs). Tbet+CXCR3+ B cells were reported to undergo autoproliferation in pwMS and to contribute to clonal differentiation from memory B cells to hypermutated plasmablasts (*46*). In our study, we did not find a separate CXCR3+ cluster, possibly because CXCR3 drives migration of ABCs to inflammation site (*47*), and we focused directly on dcLNs and not CNS. Nevertheless, we observed a similar Tbet+CD11c+ population, that based on BCR clonal overlap differentiated to plasmablasts and were only modestly depleted. The resistance of these DN B cells to anti-CD20 depletion may reflect relatively reduced surface CD20 expression compared to conventional mature B cell subsets (*48*). LN microenvironment could also provide survival niches including access to follicular stromal cells, cytokines or antigens selectively sustaining DN B cells.

During anti-CD20, we saw the DN, Tbet+CD11c+, and IFN-stimulated B cells to share clonal relationships along a naïve–memory–plasmablast differentiation trajectory rather than through canonical GC reactions. This is consistent with emerging evidence that extrafollicular B cell activation pathways, driven by TLR signaling, type I IFN, and T cell-independent stimuli, can generate class-switched and somatically hypermutated B cells and antibody-secreting cells outside the GC (*49, 50*). Such GC-independent pathways have been increasingly recognized in the context of autoimmunity and chronic viral infections, where immune stimulation may bypass the selection mechanisms of the GC reaction, thereby permitting the accumulation of potentially autoreactive or dysregulated B cell clones (*49, 50*).

High enrichment of lytic EBV host gene signatures in DN B cells in the dcLNs, before and during anti-CD20 therapy, links the B cell maturation trajectories we observed to EBV influenced B cells behind MS. EBV infected B cells mostly undergo class switching without somatic hypermutation, uncoupling canonical GC selection, as reported in a human tonsillar organoid (*51*). EBV preferentially reprograms a memory B cell subset (CD27+CD21low) toward a T-bet-high, antigen-presenting profile in patients with SLE, with the viral protein EBNA2 directly acting on the relevant gene regulatory regions (*52*). In the context of MS, Tbet+CXCR3+ B cells were the preferred B cells for EBV residency and subsequent CNS-homing in a humanized mouse model of demyelination (*53*).

After primarily infecting naïve B cells (*54*), EBV establishes latency in memory B cells with lytic reactivation occurring upon plasma cell differentiation (*55, 56*). We saw also a signature of naïve B cell lytic EBV infection during anti-CD20 therapy, which suggests local reinfection of novel naïve B cells entering the LN. This is in line with EBV acting as a continuous driver of MS pathology instead of a simple trigger effect (*57, 58*).

Of the B-T cell crosstalk, we saw a convergent LGALS9-based signaling program shared across DN, IFN-stimulated, and Tbet+CD11c+ B cell subsets, directed toward multiple CD4 and CD8 T cell populations in the dcLNs. Galectin-9 is a well-established modulator of T cell fate through TIM-3 engagement and can drive T cell death, an effect that is attenuated by co-expression of PD-1, allowing persistence of dysfunctional PD-1+TIM-3+ T cell populations (*30*). This suggests that the LGALS9 signaling program identified here could contribute to the exhausted/dysregulated phenotype of the CD4 and CD8 T cell populations engaged in this axis and may explain why CD8 T cell exhaustion was not normalized. Alternative option could be the insufficient depletion of DN B cells that keep stimulating TRM-EX and TREM-TEX cells that were the most expanded of CD8 T cells.

Clonally expanded CD8 memory T cells in pwMS retained EBV targeting during anti-CD20 in our study. While intravenous anti-CD20 (ocrelizumab) was seen to reduce EBV-specific CD8 T cell proliferative responses in peripheral blood (*59*), the overall clonal diversity of the circulating T cell repertoire has been reported to remain unaffected during anti-CD20 therapy (*60*), similar to our finding. Furthermore, rituximab-mediated lysis of EBV-infected B cells has been shown to release viral antigens that can activate memory T cells (*61*). The persistence of elevated CD8 T cell exhaustion in dcLNs of pwMS despite anti-CD20 therapy can thus reflect ongoing and inefficient attempts to control the persistent EBV infection.

Our cohort included a patient relapsing during anti-CD20 therapy, allowing for a case-based query into the differences of disease activity before treatment and during high-efficacy therapy. Relatively higher proportion of dark zone GC B cells in the dcLNs, contrasting the before treatment finding of reduced GC B cells in pwMS compared to healthy controls, suggested restored GC entry within the dcLNs. B cells were more clonal than in other patients and clonal sharing between GC dark zone B cells and various pre-treatment B cell subsets including IFN-stimulated B cells suggested that already primed B cells survived B cell depletion and re-entered GCs during a relapse.

Our study has limitations. Sample sizes for the DMF (n=2) and NTZ (n=1) treatment groups were limited. Our study cohort included a relapsing patient during ofatumumab and thus whether there are differences in the LN effects of differentially administered steady-state anti-CD20s should be resolved in larger studies. FNA sampling of dcLNs is minimally invasive and enables longitudinal sampling but it does not fully capture the spatial and architectural heterogeneity of LN microenvironments, including follicular versus interfollicular compartments. Additionally, while our EBV lytic gene signature analysis is based on a well-characterized reference dataset (*35*) and supported by the clonal lineage and T cell epitope data, novel tools for functional validation of the EBV reactivation in DN and de novo infection of naïve B cells of the dcLNs are needed. Finally, the causal relationship between surviving B cell populations and persistent CD8 T cell exhaustion, while supported by the convergence of multiple independent analyses, remains correlative and should be addressed in future mechanistic studies.

Our findings suggest a sustained, EBV-driven pathology in pwMS during anti-CD20 in the dcLNs, biologically plausible to involve CNS-directed lymphocyte priming. DN B cells, broadly implicated in autoimmunity, present with a persisting EBV lytic infection signature within the dcLNs, a finding uncaptured by investigating PBMC, and interact with exhausted T cells. EBV targeting of expanded CD8 T cell clones persists, supporting EBV as a continuous driver of MS. Our results establish a foundation for the integration of lymphoid tissue oriented and mechanism-driven approaches to understanding treatment efficacy in pwMS and highlight the dcLNs as a critical but underexplored compartment for monitoring and improving therapeutic outcomes for pwMS.

## MATERIALS AND METHODS

### Study design

We previously investigated dcLNs of newly diagnosed pwMS using ultrasound-guided FNAs (*15*). Here we follow the same approach and resample dcLNs of pwMS during immunotherapy and compare the cellular compartment of the dcLN to peripheral blood mononuclear cells (PBMC). The overview of the study design is shown in Figure 1A. A neurologist at the Department of Neurology, Helsinki University Hospital, recruited pwMS to participate in the study. All participating individuals provided written informed consent according to the principles outlined in the Declaration of Helsinki. Total 25 samples, of which 13 from newly diagnosed pwMS before treatment, 3 from healthy controls presented in the previous study (*15*), 8 follow-up samples taken from pwMS during DMTs, of which 7 from resampling of same individuals and 1 follow-up control sample were included in the comparison. Additionally, matched PBMC samples taken for a subset of participants, including 4 pwMS before treatment and 4 follow-up pwMS sampled during DMT, were analyzed. Time from baseline to follow-up sampling was a median 17 months (range 9–41; n=7). Time on DMT before follow-up sampling was a median 13.5 months (range 7-40). This study was approved by the Helsinki University Hospital ethical committee (decision number 5774/2020; research permit number 2363/2020).

### Subjects

The diagnosis of MS was confirmed by an experienced neurologist at the Helsinki University Hospital using the 2024 McDonald diagnostic criteria (*62*). The comprehensive diagnostic evaluation involved the assessment by a neuroimmunologist, cerebrospinal fluid (CSF) analysis, and a head magnetic resonance imaging study. All pwMS exhibited oligoclonal bands or elevated kappa free light chain index in the CSF at the time of diagnosis.

### Peripheral blood collection and mononuclear cell purification

Peripheral venous blood samples were collected to 10mL lithium heparin vacutainer tubes. Plasma was separated by centrifugation at 400xg 8 minutes at 4°C, and stored separately at –80°C. To isolate PBMCs, remaining whole blood was diluted with 1x PBS and poured over 15ml Ficoll-pague (Cytiva) phase and centrifuged at 400xg 30 minutes at RT, acceleration and brake off. Subsequently, the white buffy coat, that includes the PBMCs, was transferred to a new tube and was washed twice with 50 ml 1x PBS at 400g 15 minutes at 4°C. Cells were counted with LunaFX7 cell counter supplemented with acridine orange at 1:10. Cells were cryopreserved with CTL cryopreservation kit (ImmunoSpot) according to manufacturer’s instructions. Around 1x10^7 cells were stored in one vial, and vials were stored at -196°C. Prior to analysis, PBMCs were thawed and washed with CTL cryopreservation kit (ImmunoSpot) according to the manufacturer’s instructions. Cells were suspended with Mg and Ca-free PBS and transferred to the Single-Cell Analytics core unit.

EBV serology status was evaluated using anti–EBV-CA ELISA IgG and IgA kits (Microgen) according to the manufacturer’s instructions. The serology results were confirmed using a protein microarray EBV Virachip, which qualitatively tested for EBV-CA (VCA gp125 and p18), EA-D, and EBNA1. All subjects except one control (Control008) were EBV-IgG seropositive.

### Fine-needle aspirations and single-cell sequencing

Viable single cells from dcLNs of pwMS and healthy controls were isolated using ultrasound-guided FNAs and single-cell sequenced as described previously (*15*). Here, dcLN cells were collected with rinses of PBS (Mg and Ca free) at the final volume of cell suspension being 4-5ml. Suspension cells were pelleted by centrifugation at 400xg 10 minutes at 4°C. Supernatant was discarded and cells were treated with 300ul ammonium-chloride-potassium (ACK) lysis buffer (Lonza) for 5 minutes at room temperature to lyse red blood cells. Cells were resuspended with 2 ml of PBS (Mg and Ca free) and counted with LunaFX7 cell counter supplemented with acridine orange at 1:10. Subsequently, cells were pelleted by centrifugation at 400xg 8 min 4°C. After discarding the supernatant, depending on the sample cell amount, a minimum of 30 000 cells was resuspended with PBS (Mg and Ca free) and proceeded to the Single-Cell Analytics core unit.

The Institute for Molecular Medicine Finland (FIMM) Single-Cell Analytics core center at the University of Helsinki performed the 5′ RNA-seq runs and library preparation for single-cell RNA sequencing (scRNA-seq), T cell receptor (TCR)αβ, and B cell receptor (BCR) libraries using the Chromium Single Cell 5′ (v2 Chemistry Dual Index) as instructed by the manufacturer (10x Genomics).

### Single-cell RNA-seq, TCR and BCR analysis

Single-cell data were processed per sample in Seurat v5 (*63*) in R (4.2.2) using sample-specific, median absolute deviations (MAD)-based quality control thresholds. After LogNormalize normalization, selection of the 2,000 most variable features, scaling and PCA (50 PCs), dcLN and PBMC samples were integrated separately using reciprocal PCA. Clustering was performed at resolution 1.2 for dcLN and 0.8 for PBMC. Cell types were annotated with SingleR (*64*) using the Human Primary Cell Atlas and Monaco immune references, followed by manual curation based on marker genes. B cell compartments were subclustered to resolve rare populations (described in detail in the Supplementary Materials and Methods).

Changes in cell type proportions across controls and pwMS before and during anti-CD20 treatment were tested with sccomp R package (*21*), adjusting for total cell number and 10x protocol (FDR < 0.05). DEGs were identified by pseudo-bulk analysis and required a BH-adjusted p < 0.05 in at least two of DESeq2 (*26*), edgeR (*27*) and limma (*28*). B–T cell communication was inferred with CellChat v2.2.0 (*29*) (described in detail in the Supplementary Materials and Methods).

TCR and BCR repertoires were analysed with scRepertoire R package (*31*). The percentage of unique clones and normalized Shannon diversity were compared between groups by unpaired t-test. Paired TCRαβ clones from expanded (≥2 times per sample) CD8 memory T cells were matched to VDJdb with tcrdist3 (*36*), using a distance of ≤12 per chain, to predict epitope specificities. CD8 T cell exhaustion was scored with AddModuleScore using established markers (*15*), and naïve-to-memory transitions were modelled with monocle3 pseudotime (*37, 38*). BCR clonal lineages were reconstructed with Immcantation tool (*32–34*)(immcantation/suite:4.4.0), defining clones by shared IGHV/IGHJ genes, equal CDR3 length and ≥75% CDR3 similarity. Hyperexpanded clones (≥20 sequences) underwent phylogenetic inference with IgPhyML (described in detail in the Supplementary Materials and Methods).

Enrichment of EBV lytic host transcripts expressed in lymphoblastoid cell lines (LCLs) described by SoRelle et al. (*35*) in B cell subsets was tested by Fisher’s exact test (FDR < 0.05) (described in detail in the Supplementary Materials and Methods).

## List of Supplementary Materials

Supplementary Material file: Supplementary Materials and Methods, Fig S1 to S4 with legends

## Data Availability

All data produced in the present study are available upon reasonable request to the authors

## Acknowledgments

We are grateful for the patients for participation and without whom this study would not have been possible. We thank M. Rissanen, M. Supperi-Joki and T. Savisto for help with sample acquisition and processing. We thank the FIMM Single-cell Analytics unit supported by HiliFE and Biocenter Finland for scRNA services.

## Funding

This study was supported by Sigrid Juselius Foundation (to SML), Emil Aaltonen Foundation (to SML), the Finnish Medical Foundation (to SML), Competitive Research Funding of the State of Finland governed through Helsinki University Hospital grant tyh2024106 (to SML), the Medical Society of Finland (to SML) and Wilhelm and Else Stockmann Foundation (to SML).

## Author contributions

Conceptualization: J.S. and S.M.L. Methodology: Y.K-B., J.S., G.K., and S.M.L. Software: Y.K-B. Formal analysis: Y.K-B. Investigation: Y.K-B., J.S., E.A., A.S., M.S., G.K., and S.M.L. Resources: S.M.L. Data curation: Y.K-B. and J.S. Writing—original draft: Y.K-B., J.S., and S.M.L. Writing—review and editing: Y.K-B., J.S., and S.M.L. Visualization: Y.K-B. Supervision: J.S. and S.M.L. Project administration: S.M.L. Funding acquisition: S.M.L.

## Competing interests

S.M.L. reports lecture and advisory fees Alexion, Amgen, Argenx, Lundbeck, Novartis, Roche, Sanofi, Teva, UCB Pharma; travel and congress expenses Merck, Novartis, UCB Pharma. J.S. reports travel and congress expenses Merck, Celltrion. Other authors have nothing to disclose.

## Data and materials availability

All data needed to evaluate the conclusions in the paper are present in the paper or the Supplementary Materials. The Seurat-objects of the processed scRNA-seq data are available at Zenodo (*will be made available after acceptance*) with restricted access due to General Data Protection Regulation (GDPR), and data can be accessed by placing a request via Zenodo. The code to reproduce the key findings is available on GitHub (github.com/yigitbabal/MS_DMTs_cervical_lymphnodes).

## Notes

### Author Declarations

Ethics committee of Helsinki University Hospital gave ethical approval for this work (decision number 5774/2020; research permit number 2363/2020).

