## Supplementary Material file 1 for "Cervical lymph node B cell subset resists depletion and sustains lytic Epstein Barr-virus signature in patients with multiple sclerosis"

### Supplementary Materials and Methods

#### Single-cell RNA-seq, TCR and BCR analysis

Initial single cell data processing was performed individually for each sample using Seurat (v5) *(63)* in R (4.2.2). Quality control (QC) metrics, number of detected genes (nFeature_RNA), total unique molecular identifier (UMI) counts (nCount_RNA), and percentage of mitochondrial transcripts (percent_mito) were calculated per cell using Seurat. For each sample, lower bounds for nFeature_RNA and nCount_RNA were defined as the median minus three median absolute deviations (MAD) on the log10-transformed distributions, while upper bounds were set at the 99.5th percentile of the raw distributions. The mitochondrial content threshold was defined as the minimum of (median + 3×MAD) and the 95th percentile of percent_mito per sample, to provide a conservative, sample-specific cutoff. These thresholds were used to guide manual inspection of QC distributions prior to cell filtering. After quality control, each sample dataset was normalized using the Seurat LogNormalize method; the 2000 most variable features were found, the data was scaled, and linear dimensional reduction was performed using principal component analysis (PCA). Number of PCs were calculated by JackStraw method in Seurat and first 50 PCs were selected for further downstream processes. Both dcLN and PBMC samples were integrated separately with reciprocal (R)PCA integration algorithm built-in Seurat. Cell clusters and UMAP dimension reductions were performed with different range of resolution parameters (from 0.2 to 1.2) and 1.2 resolution were selected for dcLN and 0.8 resolution for PBMC. The cell types were determined by first using human primary cell atlas and Monaco immune atlas in SingleR *(64)* and then manually adjusting clusters based on gene expression profiles. Additionally, B cell compartments annotated in dcLN and PBMC were subclustered to identify rare sub-populations.

Changes in cell type proportions across control, before, and after-treatment samples from pwMS were identified using a sum-constrained beta-binomial model in the sccomp R package *(21)*. To account for batch effects across samples, the total number of cells per sample and 10x sequencing protocol were included as covariates in the model. Cell type proportions with FDR $< 0.05$ were considered significantly altered.

Differentially expressed genes (DEGs) between cell types and groups were identified using a pseudo-bulk approach. To increase the accuracy of differential gene expression analysis, a consensus of three well-known DE analysis tools (DESeq2 *(26)*, edgeR *(27)* and limma *(28)*) was applied, and genes were identified as DEGs when they have statistical significance (Benjamini-Hochberg adjusted p-value $< 0.05$) in at least two of the three tools.

For the investigation of B and T cell interactions, we performed ligand-receptor based cellular communication analysis using CellChat (v2.2.0) *(29)*. CellChat objects were generated with normalized expression matrices. After identifying ligand-receptor pairs for every cell type, we focused on B and T cell interaction by combining normalized expression values of ligand-receptors from corresponding cell types and samples.

Comparison of the TCR and BCR immune repertoires were performed by scRepertoire R package *(31)*. Percentage of unique BCR/TCR clones and normalized Shannon diversities were compared across control, before treatment and during anti-CD20 treatment taken samples from pwMS by unpaired t-test. Unique clonetypes were defined as VDJ gene segments and amino acid sequence together. Expanded TCR clones were identified as TCR clones ≥ 2 times in a sample. TCR clones belonging to clonally expanded CD8+ memory T cells were used for public VDJdb repertoire matching using tcrdist3 *(36)* to predict epitope targets. Matching was performed on paired TCRαβ clones using a tcrdist3 distance threshold of ≤12 per chain for both the alpha and beta chains, corresponding to approximately one amino acid mismatch per chain, to assign predicted epitope specificities to our TCR sequences.

Exhaustion scores were calculated for the CD8+ T cells with *AddModuleScore* function in Seurat using previously described exhaustion markers *(15)*: TIGIT, HAVCR2, PDCD1, CD244, LAG3, TCF7, and CTLA4. To identify exhausted CD8+ T cells, T cell compartments were subclustered, and marker gene expression were combined with exhaustion module scores. Naïve to memory CD8+ T cell transition was analyzed with monocle3 *(37, 38)* pseudotime ordering to investigate the dynamics of exhausted gene expression over pseudotime.

BCR clonal lineages were constructed across before treatment and during anti-CD20 treatment samples of pwMS with Immcantation tool *(32–34)*(immcantation/suite:4.4.0). Clones were defined by shared IGHV gene, IGHJ gene, and CDR3 length with ≥75% CDR3 sequence similarity (Hamming distance threshold 0.25). Clones with ≥2 sequences were considered expanded and included in lineage tree construction. Clones with ≥20 sequences were classified as hyperexpanded and were subjected to phylogenetic tree inference using IgPhyML with the GY94 codon substitution model.

EBV lytic gene enrichment analysis was performed for the B cell subsets with Fisher-exact test by using lytic host transcripts expressed in lymphoblastoid cell lines (LCLs) described by SoRelle et al. *(35)*. Statistically significant EBV lytic marker enrichments for B cell subsets were identified with FDR $< 0.05$ threshold.

### Supplementary Figures

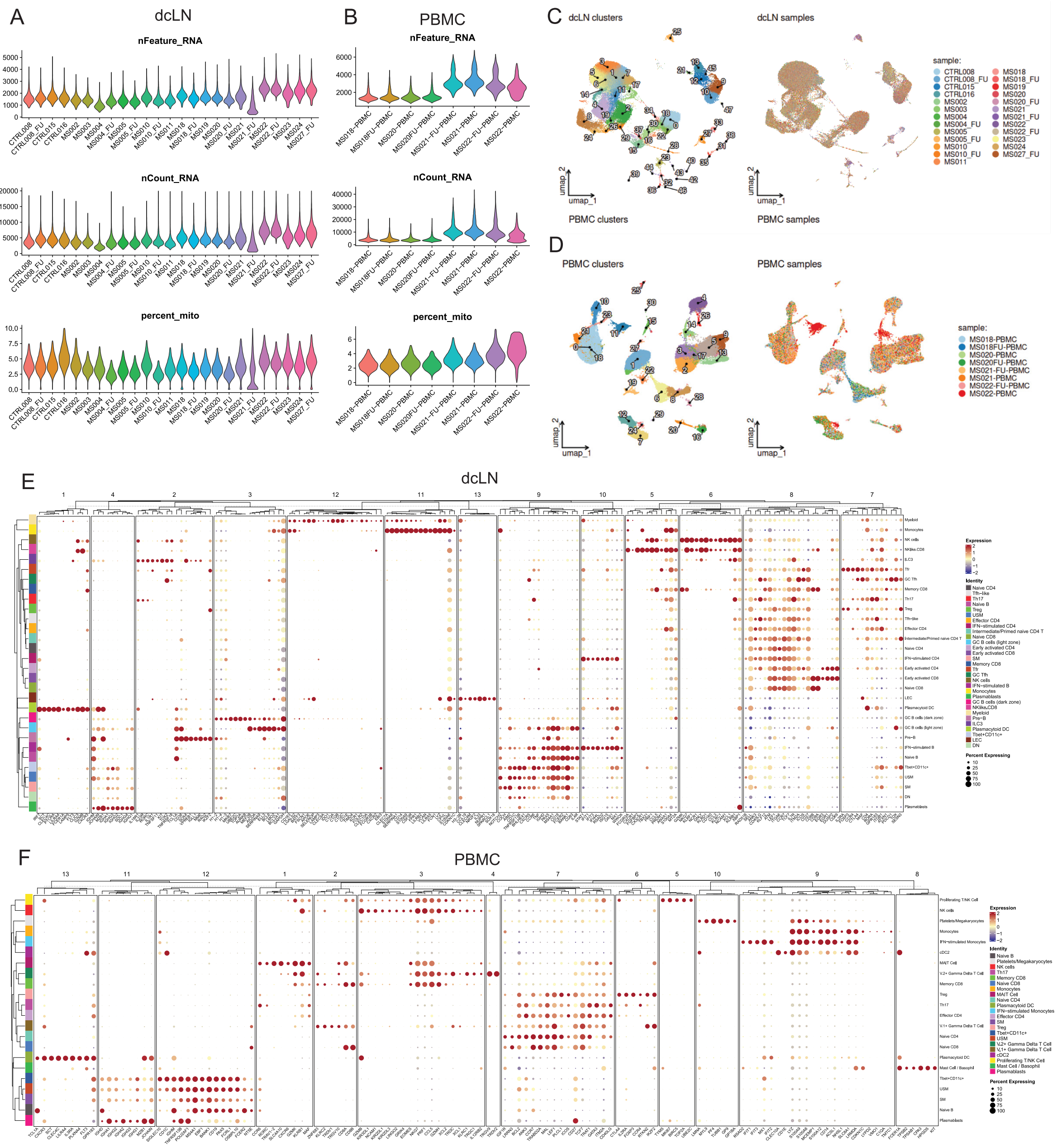


### Fig. S1: Quality control and cell type annotation of dcLN and PBMC scRNAseq data.

(**A**) Violin plots presenting quality control metrics (total number of features, total number of counts and mitochondrial gene percentage) of dcLN and (**B**) PBMC scRNA-seq. (**C**) UMAP representation of Seurat clusters and sample distribution of dcLN FNA and (**D**) PBMC samples. (**E**) Dot plots showing comprehensive marker gene expression used in cell type annotation of dcLN and (**F**) PBMC.

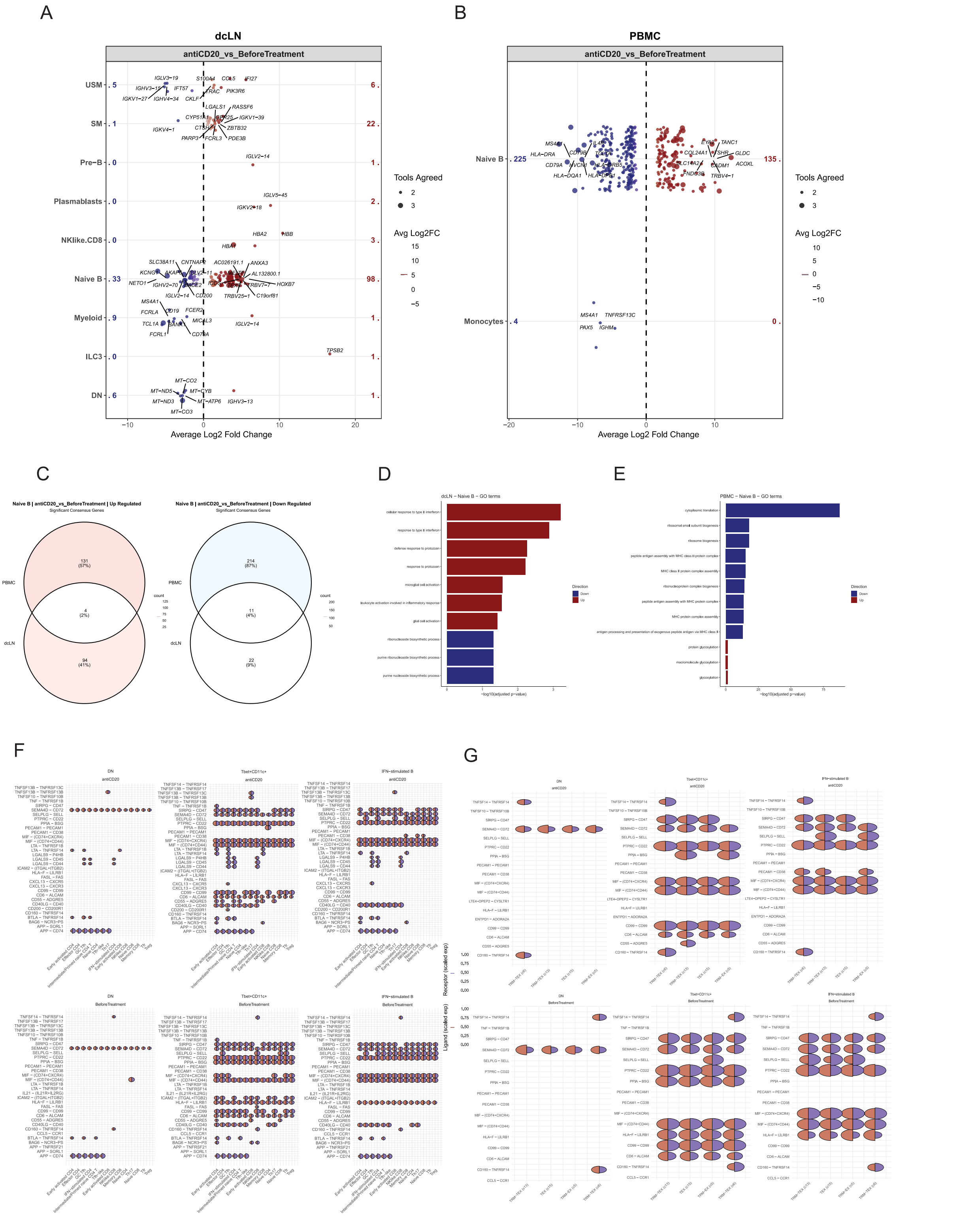


**Fig. S2: Cellular and molecular change of dcLNs and PBMC during anti-CD20 therapy.**

(**A**) Dot plots of consensus differential gene expression results in dcLNs and (**B**) circulation. DEGs for each cell type of dcLNs and PBMC in pwMS under anti-CD20 therapy compared to before treatment pwMS with pseudobulk approach. Log2FC presented in x-axis shows average significant log2FC as determined to require at least two tools coherence (DESeq2, edgeR and limma). (**C**) Venn diagram of common and unique DEGs of naïve B cells in dcLN and PBMC. (**D**) Bar plots of enriched GO terms performed for up- and down-regulated DEGs of naïve B cells in dcLNs and (**E**) PBMC. (**F**) Mushroom plots of ligand-receptor based cell-to-cell communication between T (ligand) and B (receptor) cell compartments in dcLNs before and after anti-CD20 therapy. Colors indicate the scale expression of corresponding ligand/receptor expression and size of the circle corresponds to expression percentage of ligand/receptor in given cell type. (**G**) Mushroom plots of ligand-receptor interaction between the exhausted T cell cluster and B cell subsets.

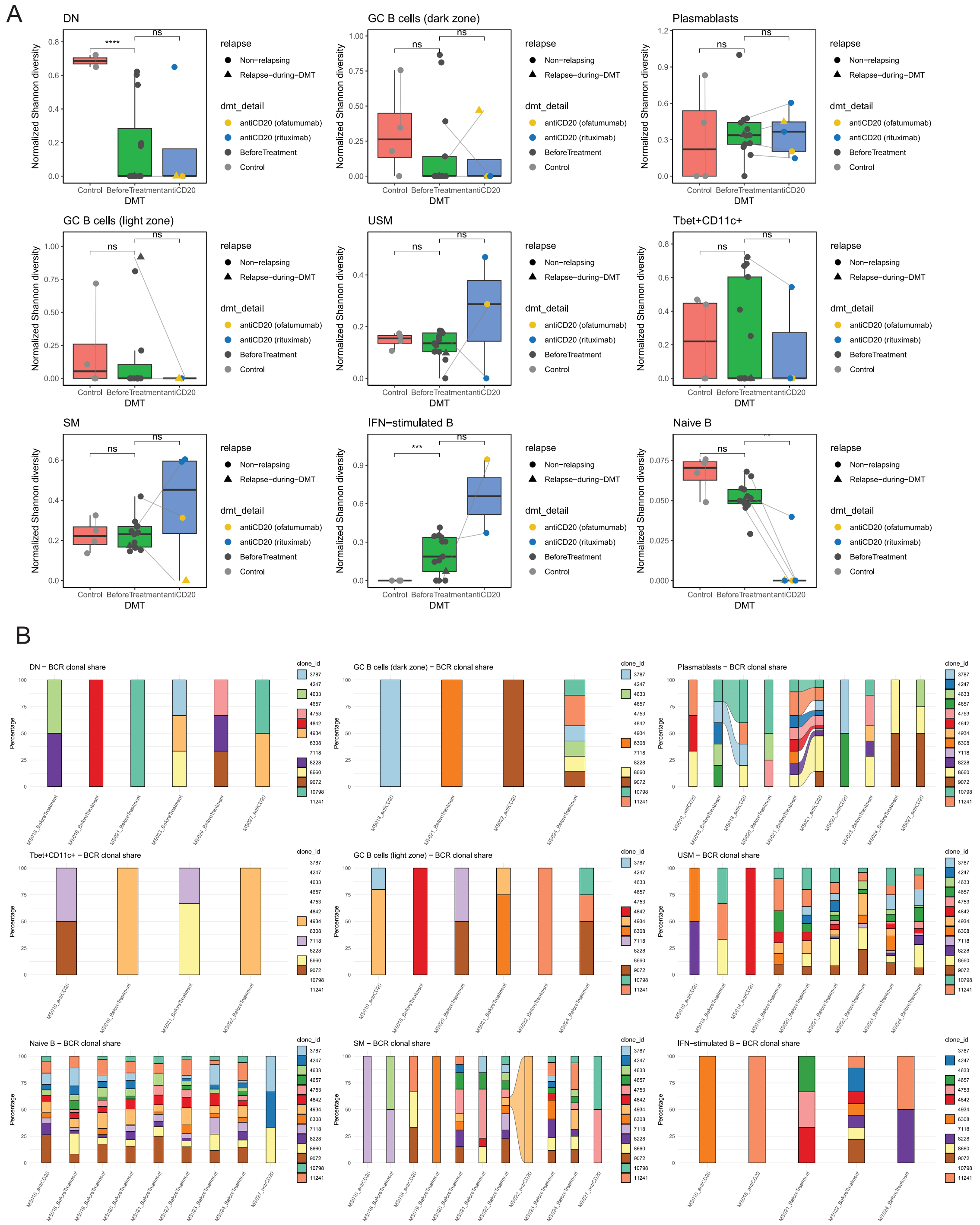

**Fig. S3: BCR clonal diversity and share in dcLNs.**

(**A**) Box plots show BCR diversity of B cell subsets in dcLNs by normalized Shannon diversity for pwMS before treatment, during anti-CD20 and for the healthy controls. (**B**) Stacked bar plot presenting BCR clonal share of B cell compartment before and after anti-CD20 therapy.

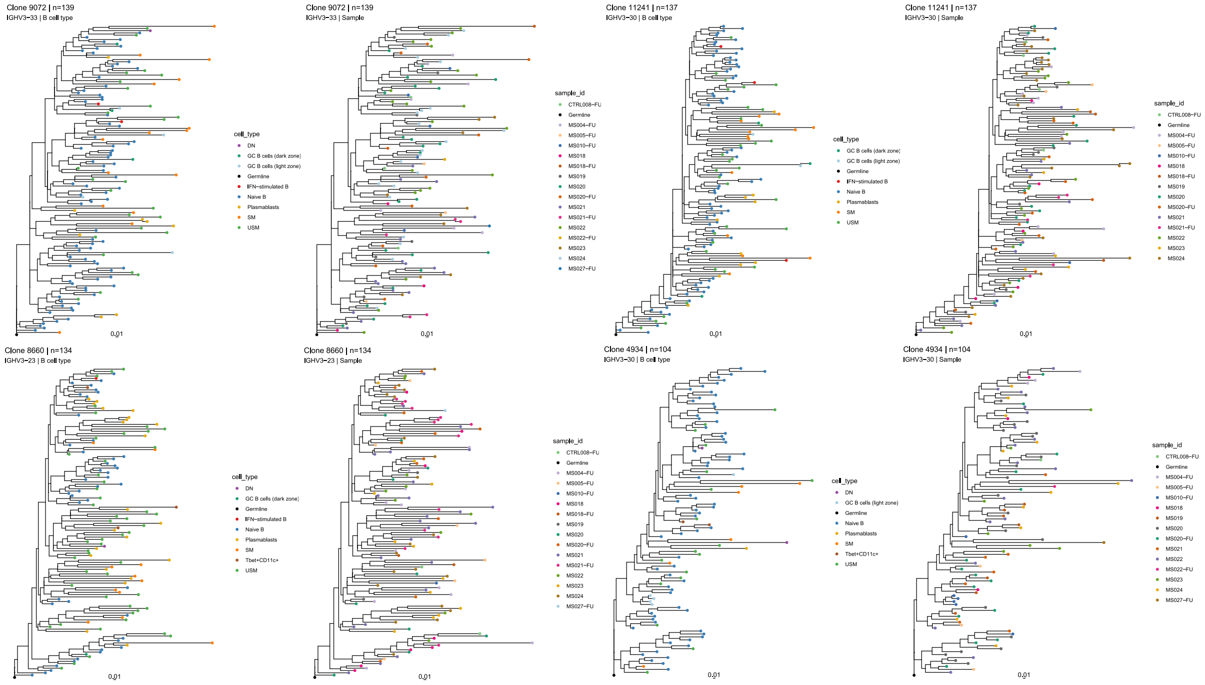


**Fig. S4: Somatic hypermutation (SHM)-aware germline trees of BCR clones in dcLNs for pwMS before treatment, anti-CD20 and healthy controls.**

Top germline trees resulted with immcantation suite showing BCR clonal share of B cell compartment. Each individual tree corresponds to particular BCR clones and length of the branches indicate SHM rates.
